# Maternal Mental Health and Engagement in Developmental Care Activities with Preterm Infants in the NICU

**DOI:** 10.1101/2022.12.09.22283171

**Authors:** Sarah E. Dubner, Maya Chan Morales, Virginia A. Marchman, Richard J. Shaw, Katherine E. Travis, Melissa Scala

## Abstract

**Objective:** **To examine associations between maternal mental health and involvement in developmental care in the NICU**.

**Study Design:** Mothers of infants born at <32 weeks gestation (n=55) were screened for anxiety, depression, and post traumatic stress disorder two weeks after admission. Mothers’ frequency, rate, and duration of developmental care activities were documented in the electronic health record. Group differences between unscreened mothers, and mothers with and without elevated screener scores and developmental care measures were assessed retrospectively.

**Results:** 35% of screened mothers scored above the cutoff for clinical concern on ≥1 measure. No significant group differences were identified for rates, frequency, or amount of overall developmental care, kangaroo care, or holding.

**Conclusion:** Maternal mental health ratings did not relate to developmental care. Maternal developmental care engagement may not indicate mental health status. Universal screening for psychological distress is required to accurately detect symptoms in mothers of hospitalized preterm infants.

## Introduction

Developmental care, which includes parent involvement in activities such as kangaroo care, positive touch, massage, and auditory exposures, for preterm infants in the neonatal intensive care unit is important to mother-infant attachment (1–6) and to infant health outcomes.(7) Mothers of preterm born infants are at elevated risk for postpartum depression, anxiety disorders, and symptoms of post traumatic stress.(8–12) These maternal mental health conditions influence mother-infant attachment,(13,14) increasing risk of maladaptive family dynamics such as vulnerable child syndrome.(15) Maternal mental health is an important factor associated with many childhood outcomes, including physical growth, childhood illness, cognitive development, and academic achievement.(3) Studies have yet to examine potential links between mental health conditions and maternal engagement in developmental care activities.

Developmental care practices in the neonatal intensive care unit have been used to assist parents in developing a healthy attachment with their critically ill infants.(15,16) Despite comprehensive and well documented developmental care programs, wide variation exists in the actual parent and infant experience of these developmental care practices.(16–18) Institutional, socio-demographic, and infant factors are known to influence developmental care implementation. In this investigation we ask whether maternal mental health contributes to the variation in developmental care practices. We hypothesized that mothers who screened positive for symptoms of psychological distress would participate in fewer developmental care activities overall. We base this hypothesis on research that depressed mothers have heightened risk of impaired infant bonding (19–22) and may have altered caregiving behaviors.(8,23–25)

## Subjects and Methods

### Participants

Participants in this retrospective study were all mothers and their infants (*n* = 135) who were born very or extremely preterm (≤ 31 6/7 weeks gestational age) between 5/1/2018 and 2/9/2020. All infants were cared for at Lucile Packard Children’s Hospital (LPCH) from admission through at least 35 weeks post menstrual age. We included infants born at other hospitals (n=19) only if they had been transferred into the LPCH neonatal intensive care unit (NICU) before 7 days of life. We chose this criteria to ensure reliable characterization of their developmental care experience for the duration of hospitalization for preterm birth. LPCH nursing education includes training for developmental care charting. Nurses routinely chart all visitation and developmental care activities (e.g., kangaroo care, swaddled holding, massage) in the electronic medical record (EMR). We excluded infants born after 2/9/2020 because of changes in hospital visitation policies that were instituted on 3/8/2020 due to the SARS-CoV2 pandemic. This cut-off date ensured that all infants experienced at least 1 month of their hospital course during the pre-pandemic period. The Stanford University Institutional Review Board approved the experimental protocol (#IRB-54650). All data were collected in the course of routine clinical care. Participants were not required to give consent for this retrospective analysis. The study was performed in accordance with the Declaration of Helsinki.

Infant and maternal characteristics were extracted from the EMR. Infant characteristics included child sex assigned at birth, gestational age at birth, gestational age at admission, and length of stay. We also tallied presence/absence of common clinical conditions and treatment which may influence receipt of developmental care, including bronchopulmonary dysplasia, necrotizing enterocolitis, intraventricular hemorrhage, and sepsis. We calculated a summary health acuity score as the sum of the presence of these four clinical conditions (0 - 3 possible score), grouping infants into those with none versus those with one or more of these conditions. Maternal characteristics included maternal age at delivery, delivery type (vaginal or cesarean section), and health insurance type (public vs. private). We previously reported that non-English speaking mothers in our NICU have lower rates of kangaroo care than English speaking mothers,(16) therefore we also collected information on maternal language preference.

### Maternal Mental Health Assessment

Mothers of infants were offered mental health services approximately 2 weeks after admission. Mothers who accepted met with a NICU clinical psychologist or social worker and completed three mental health screening measures of psychological distress. The Patient Health Questionnaire (PHQ-9) screens for depression and consists of nine prompts asking about the frequency of symptoms over the past two weeks (e.g., “Little interest or pleasure in doing things”) to which patients responded from “not at all” to “nearly every day”.(26) The possible score range is 0-27; scores >15 were considered clinically elevated. The Neuro-QoL Short Form v1.0 - Anxiety (NQOL) consists of eight prompts about unpleasant thoughts and/or feelings related to fear, helplessness, worry and hyperarousal (e.g., “I felt uneasy”) scored from 1-5 (“never” to “always”).(27) The possible range is 8-40, with > 26 considered clinically elevated. The Perinatal Post Traumatic Stress Disorder Questionnaire (PPQ) consists of 14 questions asking about reactions to their infant’s birth (e.g., “Did you have bad dreams of giving birth or of your baby’s hospital stay?”), scored from 0-4 (“not at all” to “often, for more than a month”).(28) The possible range is 0-56, with scores > 19 considered clinically elevated. Mothers who scored in the clinically elevated range on any of the questionnaires were counseled by the clinical psychologist and referred to hospital or community resources for further assessment and symptom management.

Of the 135 infant-mother dyads, 55 (41%) completed the mental health screening questionnaires. For analyses, we grouped mothers who participated in the screening into two cohorts based on whether they scored in the clinically elevated range on one or more of the assessments: clinically elevated (*n* = 19) and not-clinically elevated (*n* = 36). Non-screened mothers were those mothers who did not complete the screening (*n* = 80). Ten (18%) scored above the cutoff of the PHQ-9 depression screener, 17 (31%) scored above the cutoff on the NQOL anxiety screener, and 11 (20%) scored above the cutoff on the PTSD screener. Of the 19 scoring above the cutoff, 8 scored in the range of concern on one screen, 3 on 2 screeners, and 8 on all three screeners.

### Developmental Care

As part of routine charting, nurses documented each instance of any developmental care activities including the type (Kangaroo Care, Swaddled Holding, Touch, Massage, Music, Talking, and Singing), the approximate duration, and who was involved (mother, father, other family member, nurse, other staff member, or any combination of these). For the purposes of these analyses, we restricted our computation to only those developmental care instances when the mother participated, either alone or with staff or other family members. A summary measure of all developmental care was generated by summing all instances of any type in which the mother was reported to be present. Because kangaroo care (skin-to-skin holding) has been used as a therapeutic intervention for mood disorders in the NICU and may be later replaced by swaddled holding, we secondarily assessed these individual care practices, again only counting those instances in which the mother was present either alone or together with staff or other family member(s).

Three measures were calculated for each developmental care type:

a. <u>Frequency</u> = sum of the instances of developmental care/total length of stay (instances/day)
b. <u>Rate</u> = total time performing developmental care activity/total length of stay (minutes/day)
c. <u>Duration</u> = total time performing developmental care activity/sum of instances of developmental care (minutes/instance)

### Statistical approach

Statistical analyses were conducted using the Statistical Package for the Social Sciences (SPSS version 28.0, IBM Corp., 2021). Two-sided statistical significance was set at p<0.05. Based on normality, we used parametric t-tests or ANOVAs for assessing group differences in continuous variables and Chi-square tests for group differences in dichotomous variables. Post-hoc tests evaluated sub-group differences applying Tukey correction.

## Results

**Table 1** shows the demographic and clinical characteristics of the infants and mothers. The distribution of female to male infants was similar across all three groups of infants (χ^2^ = .63, *p* = .73) and there were no group differences in the age of admission to the NICU, *F*(2, 132) = 0.30, *p* = .75. However, there was a significant group difference in gestational age (GA) at birth, *F*(2, 134) = 5.9, *p* < .001, such that the infants of the non-screened mothers were significantly older at birth than those in the screened non-clinically elevated group (*p* < .03) and the screened clinically-elevated group (*p* < .004). Infants of mothers in the two screened groups were not significantly different in GA at birth, *t*(53) = 1.21, *p* = 0.23. In addition, length of stay was significantly different across the three groups, *F*(2,132) = 5.6, *p* = .005, with those infants in the screened clinically-elevated group staying in the hospital significantly longer than infants in the non-screened group (*p* < .01). While there was a trend for a higher proportion of infants with one or more medical complications in the screened groups, this difference did not achieve significance (*p* = .13). There were no group differences in maternal age (*p* = .29), nor in the frequency of cesarean mode of delivery (*p* = 0.6), public (vs. private) insurance (*p =* .85), or non-English preferred language (*p* = .99).

**Table 1.**
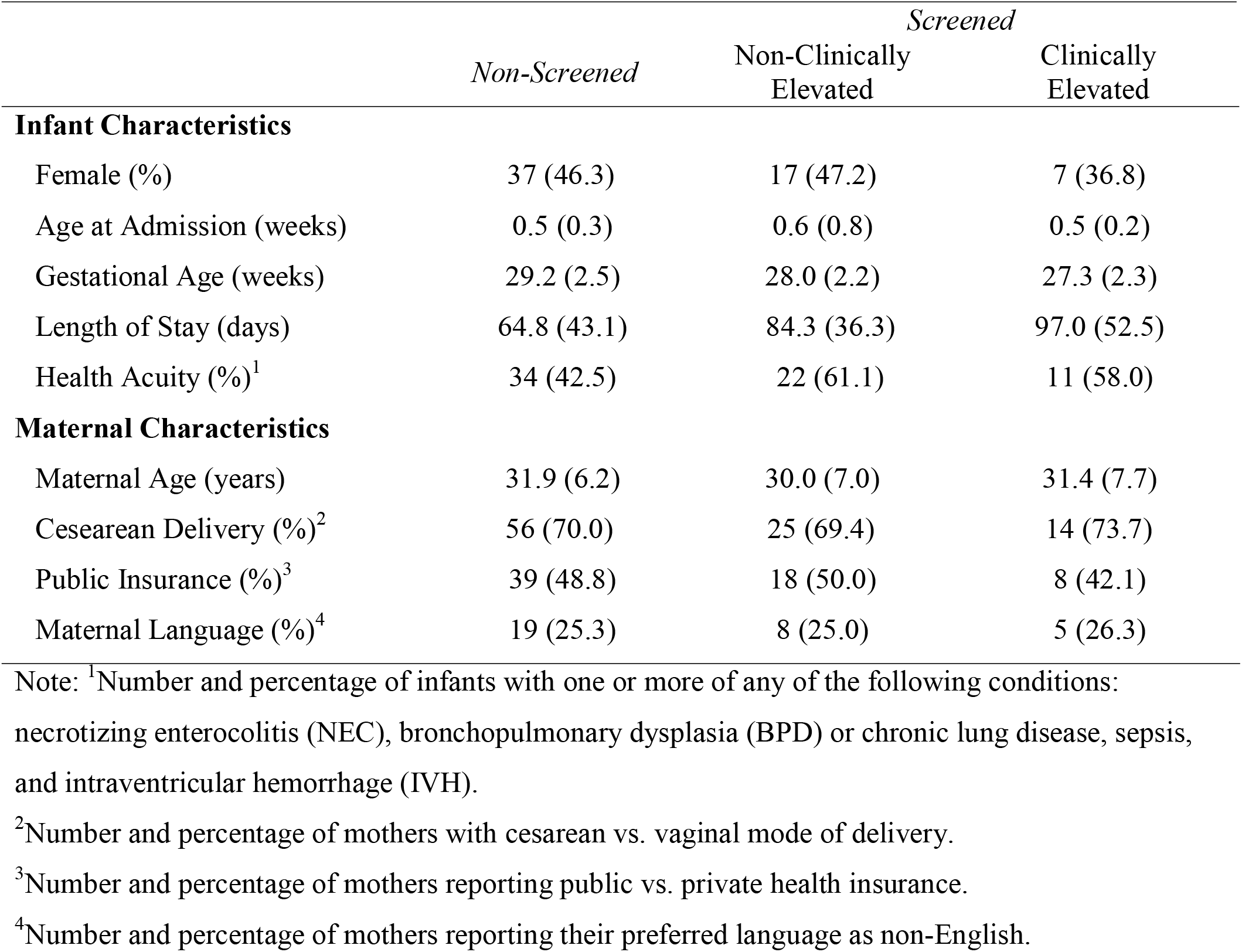
Characteristics (M (SD) or n (%) of Non-Screened (*n* = 80) and Screened (*n* = 55) infants and mothers, with screened mothers further classified into Non-Clinically Elevated (*n* = 36) and Clinically Elevated (*n* = 19) groups

**Table 2** shows that mothers in all groups engaged in developmental care activities just over once per day, on average, with no significant group differences in the frequency of developmental care activities, *F*(2, 134) = 0.15, *p* = 0.86. The pattern of similar levels of developmental care activities did not change even when controlling for length of stay and GA at birth, *F*(3, 130) = 0.27, *p* = 0.76. Developmental activities resulted in about 60 minutes/day of developmental care for all infants, on average, with each developmental care instance lasting about 50 minutes/instance. Both the rate and duration of all maternal developmental care activities did not differ significantly across the three groups (*p* > 0.72), even when controlling for GA and length of stay (all *p* > 0.71). Because we were specifically interested in differences between those mothers who scored as elevated on one or more of the screening measures versus those who did not, post-hoc comparisons explored group differences just among mothers in those two groups. Again, there were no significant differences in any measure even after controlling GA and length of stay (all *p* > 0.57).

**Table 2.** Frequency, rate, and duration of three types of Developmental Care (M (SD)) activities (all, kangaroo care, swaddled holding) performed by mothers who did not receive mental health screening (Non-Screened; *n* = 80) versus two groups of mothers who did receive mental health screening (non-clinically elevated; *n* = 36; clinically-elevated; *n* = 19)

|  | *Non-Screened* | *Screened* |  |
| --- | --- | --- | --- |
|  |  | Non-Clinically Elevated | Clinically Elevated |
| All Developmental Care |  |  |  |
| Frequency | 1.2 (0.8) | 1.2 (0.8) | 1.3 (0.5) |
| Rate | 61.5 (43.2) | 61.6 (49.6) | 66.7 (28.0) |
| Duration | 57.9 (10.2) | 57.0 (10.9) | 58.5 (9.5) |
| Kangaroo Care |  |  |  |
| Frequency | 0.2 (0.2) | 0.2 (0.2) | 0.2 (0.1) |
| Rate | 17.6 (18.5) | 14.0 (12.6) | 17.1 (10.3) |
| Duration | 69.6 (14.9) | 71.2 (15.7) | 77.4 (21.2) |
| Swaddled Holding |  |  |  |
| Frequency | 0.5 (0.4) | 0.5 (0.5) | 0.5 (0.3) |
| Rate | 33.1 (27.2) | 35.3 (36.3) | 34.5 (17.7) |
| Duration | 63.1 (11.9) | 65.6 (10.2) | 67.3 (12.5) |

Note: Frequency = Total number of instances divided by number of days of hospitalization; Rate = Total minutes divided by number of days of hospitalizations; Duration = Total minutes divided by total number of instances

While there were no significant differences across all developmental care activities, it is possible that maternal mental health status may have moderated participation in specific activities, such as skin-to-skin (kangaroo) care and swaddled holding. As shown in **Table 2**, the frequency, rate and duration of kangaroo care and swaddled holding activities was similar across the groups with no statistically significant group differences in any measure (all *p* > .27). Again, post-hoc comparisons evaluated differences just between those mothers with clinically-elevated scores vs. those screened mothers with non-clinically-elevated scores. No significant differences were found (all *p* > .37). Finally, we evaluated group differences in rates of these specific developmental care activities, controlling for other factors that may have affected rates of developmental care, including GA and length of stay. The addition of the covariates did not change the pattern of the results.

## Discussion

In this investigation of mothers of infants born very or extremely preterm we found that the amount of developmental care activities performed by mothers did not differ between mothers with or without clinically elevated self report of symptoms of depression, anxiety, or PTSD symptoms. No differences were found in the rate (total minutes of developmental care activities per day), nor in the frequency or duration of all developmental care activities and of kangaroo care or holding practices alone. Furthermore, we found no differences in the amounts of developmental care activities performed by mothers who had or had not accepted to participate in mental health screening.

These findings were contrary to our initial hypothesis that mothers with clinically significant mental health concerns would engage in less developmental care. While there is an established relationship between maternal mental health and mother-infant interactions and engagement in preterm mother-infant dyads post discharge,(13,19,20,22) maternal mental health may not have the same salience in the NICU environment where parents benefit from more intensive structure and support for parenting activities. Parenting activities in the NICU may be more tightly guided by unit policies, infant health and support needs, and infant maturity levels. In addition, parents receive daily prompting from the healthcare team to engage in specific practices so may be less able to adjust activity levels depending on mental health. Moreover, all mothers who scored above clinical thresholds were offered mental health services which likely included encouragement to participate in developmental care activities, making these activities a conscious therapeutic act rather than part of routine care. Note that all of these factors are specific to the NICU experience and therefore our results may not be generalizable to other contexts, such as the home environment. Future research should also investigate possible long term and cumulative sequelae of maternal psychological distress during the neonatal period on maternal engagement after discharge to home.

Kangaroo care specifically has been associated with improved maternal mood (29–31) and has been studied as a mental health intervention.(32) In this sample, mothers with clinically elevated symptomatology did no more kangaroo care than those without clinically elevated scores. Finally, other factors may be more powerful drivers of developmental care in the NICU, overwhelming the impact of symptoms of maternal psychological distress. For instance, in our previous research we found that lower socioeconomic status or being of lower English proficiency was significantly associated with reduced amounts of developmental care activity among parents.(16) Our findings suggest that factors such as developmental care policies, psychological support services for parents and equal access to developmental care education and engagement may provide more important modifiable avenues to improve rates of developmental care.

The present study provides novel evidence that elevated scores on screening measures of maternal psychological distress may not significantly contribute to variation in the total rates, frequency, and duration of developmental care experienced by preterm infants in the NICU. Clinicians and staff caring for preterm infants and their families should take caution - maternal engagement in developmental care activities with their infants is not a marker for maternal mood and well being. One might imagine that a particularly anxious mother might visit her infant more or that a mother with many depressive symptoms might engage less, but these would be misattributions. Our analysis suggests that maternal participation in developmental care activities does not indicate presence or absence of significant maternal distress. This analysis contributes to the body of evidence for instituting universal screening for mood disorders among mothers of preterm born infants with appropriate referral and management supports.

The study had limitations. The data were extracted and analyzed from EMRs and clinical psychologists who administered the screening measures and thus may capture inconsistencies in reporting from clinical staff. The sample was not large enough to delve into the timing of developmental care activities or to examine developmental activities before and after screening. The study did not include information about the quality of interactions which may be an even more important indicator of parent-infant attachment and the quality of caregiving behaviors.(33–35) There is no follow up data on the initial screening tools to identify the trajectory of symptoms of psychological distress and whether improvement in symptoms was associated with developmental care activities. Our measures were self report questionnaires and we did not have clinician evaluation with medical diagnoses. Finally, there is limited information on mothers who did not complete screening measures, limiting generalizability of our results. For instance, some non-screened mothers may have declined because they were already receiving mental health services. All of these limitations offer opportunities for further research regarding the interaction between parental mood and infant caregiving in the NICU.

In summary, there were no significant relationships between clinically elevated scores on maternal mental health screener and rates, frequency, and duration of developmental care activities. It is heartening to know that perinatal maternal mental health may not be an important factor affecting engagement in NICU developmental care, especially considering the trauma and stress of preterm birth. This finding also suggests that parental engagement in developmental care activities may not serve as an appropriate predictor of mental health. While maternal mental health variables may not contribute significantly to variation in developmental care activities, high rates of parental psychological distress exist in the NICU family population and negative associations have been well documented between poor parental mental health and future neurodevelopmental outcomes in NICU infants.(3,8–10) As such, perinatal maternal mood screeners should be implemented in infant healthcare environments and longitudinally following NICU discharge, to gauge parental mental health in the acute setting and over time. Consideration should be given to serial screening, including after discharge as NICU parent mental health is a dynamic condition. Further research should be conducted to determine how engagement in NICU developmental care practices may affect parenting behaviors, including parent-infant dyad engagement, after hospital discharge.

## Supporting information

Strobe_Checklist

## Data Availability

Deidentified Data will be made available upon reasonable request to the authors.

## Additional Information

### Conflict of Interest

The authors have no conflicts of interest to disclose.

#### Ethics approval and consent to participate

The Stanford University Institutional Review Board approved the experimental protocol (#IRB-54650). All data were collected in the course of routine clinical care. Participants were not required to give consent for this retrospective analysis. The study was performed in accordance with the Declaration of Helsinki.

### Availability of Data and Materials

Deidentified Data will be made available upon reasonable request.

### Author Contributions

Sarah E. Dubner contributed to data acquisition and data interpretation, and drafted the manuscript.

Maya Chan Morales performed initial data analyses and drafted the manuscript.

Virginia A. Marchman performed data analysis and data interpretation, and revised the manuscript for important intellectual content.

Richard Shaw contributed to conception and design of the study, data acquisition, and reviewed the manuscript critically for important intellectual content.

Katherine E. Travis contributed to study design, data acquisition, data interpretation, and reviewed the manuscript for critically important intellectual content.

Melissa Scala conceputalized the study, contributed to interpretation of the data, and reviewed the manuscript for critically important intellectual content.

All authors approve the final version of the manuscript and agree to be accountable for all aspects of the work.

## Acknowledgments

The authors would like to thank the NICU nurses, clinical psychologists, and clinical research coordinator, Molly Fradin Lazarus, for their contributions to data collection. We thank the parents and families of the LPCH NICU for their generous participation in this study.

